# Public health impact of better vehicle safety standards in Mexico

**DOI:** 10.64898/2026.04.28.26351923

**Authors:** Francisco R. Mojarro, Carolina Pérez-Ferrer, Husam Muslim, Sergio Bautista-Arredondo, Stephan Brodziak, Sandra Avalos-Álvarez, Nancy Izquierdo-Gutierrez, Ariadna Juárez-Rueda, Tonatiuh Barrientos-Gutierrez, Jacobo Antona-Makoshi

## Abstract

**Background:** Implementing proven vehicle safety standards recommended by the UN World Forum for Harmonization of Vehicle Regulations is among the most cost-effective strategies to reduce road traffic deaths. In 2022, Mexico approved updated vehicle safety standards, including side pole testing, electronic stability control, seatbelts, airbags, side structures, and anchorage child restraint systems. However, pedestrian protection and advanced driver-assistance technologies, such as autonomous emergency braking systems (AEBS), were excluded. These exclusions are critical, given that more than half of road traffic deaths involve vulnerable road users. Local evidence on the expected benefits of implementing comprehensive vehicle safety standards is needed to guide policy decision-making.

**Objective:** To estimate the potential public health impact of increasing the availability of recommended vehicle safety technologies in Mexico.

**Methods:** We conducted a comparative risk assessment analysis to estimate the impact of improving vehicle safety standards on road traffic deaths, injuries, and disability-adjusted life years. Counterfactual analyses were defined using traffic statistics for 2019 as baseline, relative risk estimates associated with each safety technology, and technology penetration within Mexico’s vehicle fleet. Three scenarios were modeled: (1) full implementation of Mexico’s 2022 standards; (2) addition of crashworthiness, AEBS, and motorcycle ABS/ESC; and (3) inclusion of expanded AEBS crash configurations, lane departure warning (LDW), and lane keeping assistance (LKA) systems.

**Results:** Scenario 1 reduced deaths by 18%, injuries by 16%, and DALYs by 18%, with the greatest benefits for car occupants. Scenario 2 reduced deaths by 29%, injuries by 27%, and DALYs by 30%, benefiting motorcyclists and pedestrians the most. Scenario 3 reduced deaths, injuries, and DALYs by 41%, 38%, and 41%, respectively, benefiting car occupants and motorcyclists.

**Conclusions:** Current vehicle safety standards in Mexico are expected to reduce deaths, injuries, and disabilities, yet existing guidelines focus largely on protecting car occupants. Mexico should strive to update and strengthen its current legislation by adding technologies that protect vulnerable road users, such as pedestrians and cyclists, and to focus on technologies for motorcycle users to further reduce the burden of road traffic injuries.

## Introduction

Road traffic injuries (RTI) are a major global public health issue, accounting for over 1.19 million deaths per year [1]. Young people aged 5-29 and vulnerable road users (pedestrians, cyclists, and motorcyclists] are disproportionately affected, with the burden being particularly high in low- and middle-income countries [1–3]. In response to this problem, the United Nations launched the Global Plan for the Decade of Action for Road Safety, which aims to reduce road traffic deaths by at least 50% by 2030 [4–7]. The plan outlines strategies designed to lower the risk of these injuries and enhance the safety of all road users, promoting a comprehensive and resilient approach to road safety worldwide [4,6–8].

One of the five pillars of this global strategy focuses on vehicle safety. The World Health Organization (WHO) indicates that the global vehicle population increased by 160% from 2011 to 2020 [1]; additionally, the number of motorcycles alone has tripled from 2011 to 2021, with an increase of 175% [1,9]. The United Nations (UN) has identified ten essential vehicle safety technologies (VST) like anti-lock braking system (ABS), electronic stability control (ESC), occupant resistance (anchorages for child restraint systems, the use of child restraint systems, and seatbelts), frontal and side airbags, side door beams, side structure beams, side structure and padding, and vehicle front-end design for pedestrian protection (crashworthiness]—that, if widely adopted and enforced, could substantially enhance road safety outcomes [10–12].

Governmental endorsement of vehicle safety legislation through the World Forum for Harmonization of Vehicle Regulations (WP.29) and the implementation of New Car Assessment Program (NCAP) evaluations are essential for advancing the adoption of vehicle safety technologies [1,7,13]. The effectiveness of these technologies in reducing road injuries is well documented [14,15]; however, progress remains hindered by limited legislation, inadequate enforcement, and weak industry support. Encouragingly, regional initiatives such as Latin NCAP and advocacy groups have driven progress in Latin America by evaluating safety standards and promoting legislative reforms [1,6].

Mexico had over 126 million inhabitants in 2020 [16,17] and more than 53 million registered vehicles in 2021 [1], equivalent to a rate of 41,920 vehicles per 100,000 population [1,18]. In 2023, there were approximately 17,000 road traffic deaths, a rate of 12.4 deaths per 100,000 people [19]. Vulnerable road users were disproportionately affected, accounting for approximately 65% of all road traffic deaths [1]; specifically, motorcyclists, with a an increase of 52.3% since 2018 [19]. Mexico is not on course to achieve the Decade of Action for Road Safety targets for 2030. In 2022, the country updated its vehicle safety legislation, specifically the NOM-194 and the General Law on Mobility and Road Safety [1,20,21]. Legislation now requires a core set of vehicle safety technologies in all new vehicles; however, crucial technologies recommended by the WP.29 and WHO, such as pedestrian protection and automatic emergency braking (AEBS), remain optional. Previous studies have shown that these technologies are effective in averting 4 to 8% of road traffic deaths and are key to protecting the most vulnerable road users [11,15]. A prior study by Bhalla et al. estimated that 27.6% of road traffic deaths could be averted in Mexico if 100% of vehicles had nine safety standards [10]. The inclusion of key WP.29 technologies could represent a significant step towards lowering the burden of road traffic injuries, particularly for pedestrians, cyclists, and motorcycle users.

The overall aim of this study was to quantify the public health impact of full compliance with approved vehicle safety technologies stated in the 2022 legislation (NOM-194), an expansion to a subset of already available technologies focused on protecting vulnerable road users, and a second set of new, emerging vehicle safety technologies in Mexico. Our study extends the evidence available for Mexico by: (1) improving model estimates by using updated and country-specific mortality and population data, as well as measured data on the penetration of vehicle safety technologies; and (2) analyzing a wider set of safety technologies, including motorcycle helmets and other safety standards for motorcycles, child restraint systems, and emerging technologies.

## Methods

We implemented a comparative risk assessment analysis to estimate the number of deaths, injuries, and disability-adjusted life years (DALYs) that would be averted if all vehicles and road users in Mexico adopted selected vehicle safety technologies and the use of protective devices. We modeled three scenarios and compared them to the baseline year of 2019 [11,15]. This year was selected because Mexican regulations changed in 2022 and to avoid shocks in mortality during the COVID-19 years. The scenarios are described in detail in Table 1. Briefly, Scenario 1 considers full compliance with Mexico’s 2022 vehicle safety regulations, Scenario 2 adds four additional technologies as recommended by WP. 29 (AEBS for rear-end crashes, design for pedestrian protection, and motorcycle ABS/ESC), and Scenario 3 adds emerging technologies, specifically advanced driver assistance systems (ADAS).

**Table 1.**
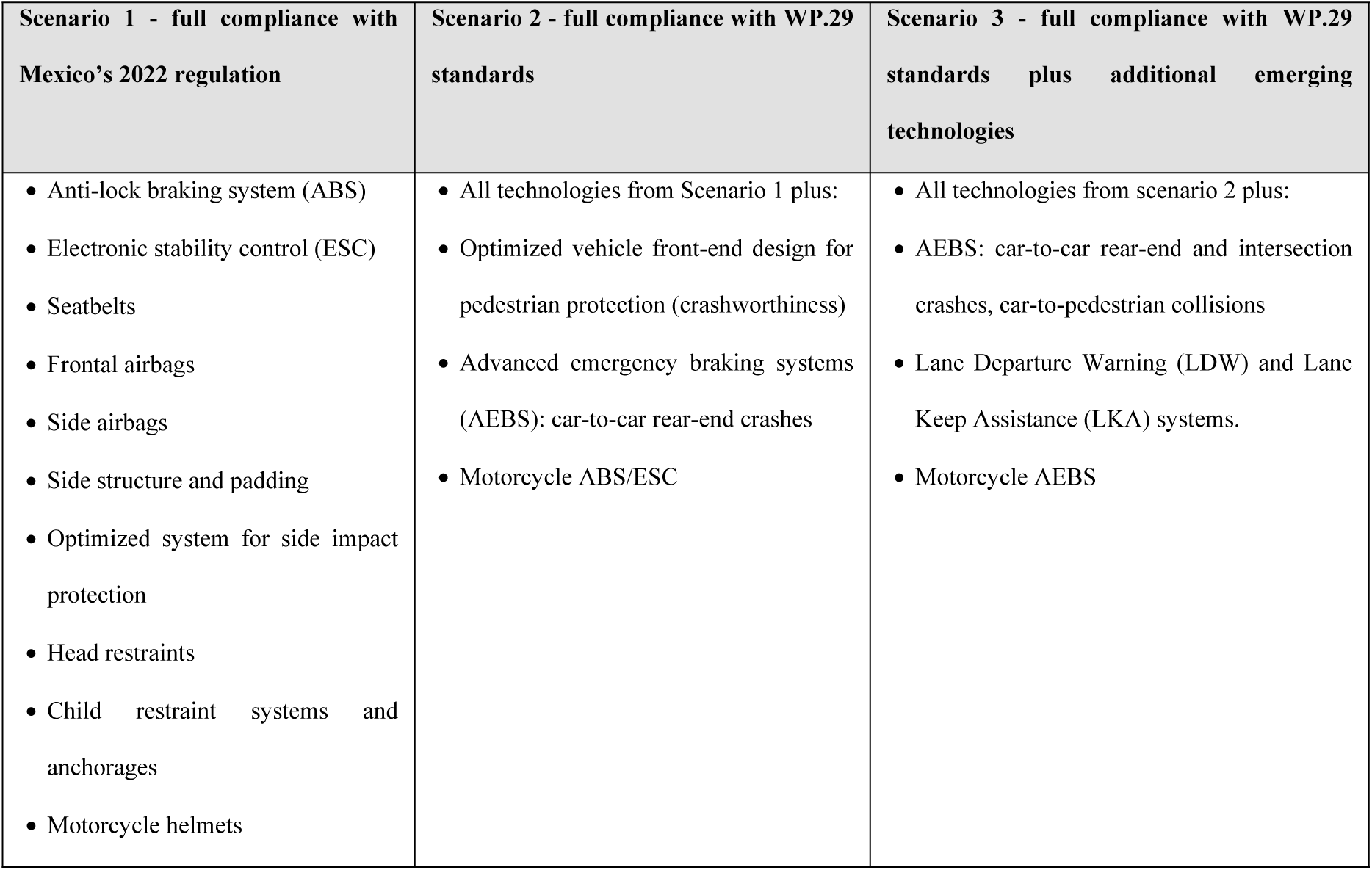
Scenarios modeled in the comparative risk assessment.

The methodology comprises four steps, as shown in Figure 1. Step 1: Data on road traffic deaths, injuries, and population distribution were collected. We also obtained data on the penetration and use of vehicle safety technologies in the 2019 vehicle fleet. Furthermore, we reviewed the literature to identify estimates of the effectiveness of each technology in preventing road traffic injuries and deaths. Step 2: We estimated the country-level burden of disease based on annual road traffic deaths and injuries in 2019 (baseline year). Step 3: We applied comparative risk assessment (CRA) to estimate the potential reduction in deaths, injuries, and DALYs associated with the full implementation of the three scenarios versus 2019. Step 4: We conducted a sensitivity analysis to test the model assumptions. (Fig **1**)

**Fig 1.**
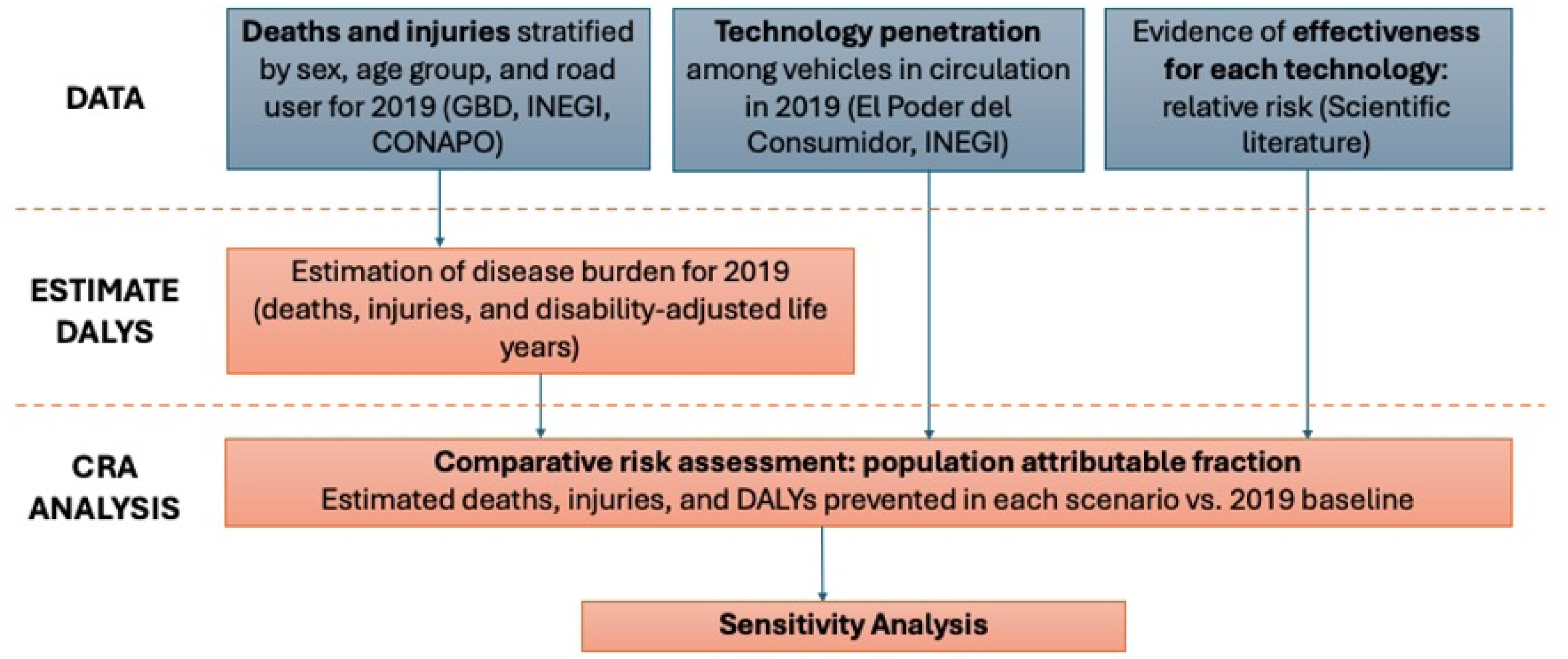
Methodology steps

### Data collection

#### Road traffic mortality and injury data

We used vital statistics data for mortality reported by the National Institute of Statistics and Geography (INEGI) [22] from January to December 2019. Mortality data were obtained from administrative death records collected through death certificates and civil registry records. This information is processed and validated annually in conjunction with the Ministry of Health (see S2 Table ***4*** for ICD codes included). We redistributed ill-defined and partially defined ICD-10 codes to account for underestimation of road traffic deaths and misclassification of road users [23,24]. The redistribution and imputation processes were applied to the following codes: V99, X59, Y31, Y32, Y33, Y34, Y85, Y85.9, Y87.2, Y89.9, V87.0-V87.9, V88.0-V88.9, and V89.0-V89.3 [25]. The detailed description of the redistribution and imputation process is available elsewhere [24,26,27]. Mortality data were stratified by sex, 5-year age group, and road user (pedestrian, bicyclist, motorized two-wheeler, car, bus, truck, and other).

Information on road traffic injuries was obtained from the Global Burden of Disease (GBD) 2019 study, which provides standardized data on non-fatal injuries disaggregated by age, sex, and road users for each country [15,26]. Official data on population’s life expectancy and mid-year total population estimates for 2019, stratified by sex and 5-year age groups, were obtained from Mexico’s National Population Council (CONAPO) [28,29].

#### Estimates of Technology Penetration and use

Vehicle safety technology penetration for 2019 was estimated using the technology prevalence among new vehicles by brand and model, vehicle sales by brand and model since 1994 [18], and an average vehicle lifespan of 26 years [30]. Data on vehicle safety technology penetration among new vehicles was obtained from “El Poder del Consumidor” (EPC), a civil society organization that advocates for better vehicle safety standards. EPC has been collecting information on vehicle safety technologies since 2015 from the top-selling brands/models in Mexico based on vehicle technical data sheets provided by vehicle makers. Their datasets include information on ABS, airbags (frontal, lateral (body/head), knees, and posterior), number of airbags, AEB, ESC, anchorages for child restraint systems, and seat belts. In 2019, EPC evaluated 96 vehicle models, which represented approximately 70% (967,089 vehicles) of all vehicles sold that year in Mexico.

To estimate vehicle safety penetration among vehicles in circulation in 2019, we first extrapolated the prevalence of each technology among new cars sold from 1994 to 2019 based on EPC data from 2016 and 2023 and modeled missing years using the typical adoption S curve [30,31]. Subsequently, using vehicle sales data for each year since 1994 and attrition rates based on Huo [30,31], we estimated the number of older vehicles still in circulation in 2019 and the proportion of vehicles equipped with each technology. The baseline penetration of each technology is presented in S3 Table ***5***. Data on seatbelt, motorcycle helmet, and child restraint system use were obtained from World Health Organization (WHO) reports and studies on road safety in Mexico [1,32–43]. Vehicle sales data were sourced from INEGI [44].

#### Vehicle safety technology effectiveness

We reviewed the literature to obtain the risk ratio (RRs) of injuries and deaths for the population exposed to vehicles with and without technologies, prioritizing evidence from systematic literature reviews. S3 Table ***5*** summarizes the RRs of death and injury for each technology or group of technologies and their references.

The effect of different vehicle safety technologies on injuries and deaths is not independent. For example, seatbelts, airbags, and ABS activate simultaneously in the case of a frontal crash and protect car occupants. Therefore, for the overall impact, we separated the global benefits of technologies by road user. For pedestrians, the model included RRs for vehicle front-end design and ABS/ESC. For motorcycles, motorcycle ABS/ESC/AEBS and helmets and for car occupants, we used the annual reduction in occupant injury or mortality risk due to *vehicle design improvements* from the National Highway Traffic Safety Administration (see S3 Table ***5*** for more details) [45], assuming that Mexico’s fleet had safety characteristics similar to those in the USA from 1980 to 2000. [7] For scenario 2 the calculations for car occupants were done in sequence, first we applied the reduction by AEBS for rear-end car-to-car collisions, then we considered the output from AEBS as an input and applied the *vehicle design improvements* reduction. Similarly, in scenario 3, calculations were done in sequence, we first estimated reduction by technology and road user group i.e. LDW/LKA (affects car occupants only), AEBS car-pedestrian (affects pedestrians only). The output of this first step was used as an input to estimate reductions from AEBS intersections (affects only car occupants), then this output used to estimate reduction from AEBS rear-end-crashes (affects only car occupants), finally this output used to estimate reductions from *vehicle design improvements*.

### Estimating the country-level health burden in 2019

DALYs were calculated using injury and mortality data, using a burden calculator developed by Bhalla and Harrison [10,46]. This tool requires population data disaggregated by sex and age group. DALYs indicate the number of years lost due to disability or premature death, with the WHO defining a DALY as one year of full health loss. In this context, DALYs were calculated as the sum of Years of Life Lost (YLL) from premature mortality due to road traffic injuries and Years Lived with Disability (YLD) associated with these injuries. YLL were obtained by subtracting the actual age at death from the expected life expectancy for Mexico in 2019, 71.8 years for men and 78 for women [47], while YLDs were calculated using the prevalence of road traffic injuries and their respective disability weights [28].

### Comparative risk assessment

We conducted a comparative risk assessment to estimate the potential changes in road traffic deaths, injuries, and DALYs if the penetration of vehicle safety technologies changed to an alternative (counterfactual) distribution. To estimate the impact of the counterfactual, we used the population attributable fraction (PAF). The PAF (Equation 1) reflects the expected proportional decrease in deaths, injuries, or DALYs if exposure to a risk factor (or a protective factor, such as a vehicle safety technology) changes to a counterfactual distribution.

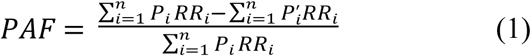

Where *P* is the proportion of the population in the *i-*th exposure category in 2019 (e.g., 33% ABS penetration), *P’* is the proportion of the population in the *i-*th exposure category in a counterfactual scenario (e.g. 100% ABS penetration), *RR* is the relative risk of death or injury for the *i-*th exposure category (e.g. relative risk of death for a car occupant involved in a crash with vs without ABS). The number of exposure categories is denoted by *n* (e.g. female, aged 30 to 34, car occupant). The PAF was computed for each technology separately and for a combination of technologies. The counterfactual (*P’*) was set at 100% for all technologies and scenarios.

### Sensitivity analysis

To evaluate the robustness of the model estimates, we conducted a sensitivity analysis considering the uncertainty associated with several key inputs, including road traffic deaths and injuries, the relative risk reduction associated with each safety technology, and estimates of technology penetration and usage. Many of these parameters were derived from multiple data sources and study designs. In particular, the effectiveness of vehicle safety technologies reported in the literature varies across evaluation methods such as field studies, experimental testing, and analyses of real-world crash data.

To account for this uncertainty, we used the mean relative risk values from the most robust evaluations as the main estimates and explored the impact of alternative assumptions by applying the minimum and maximum values reported in the literature. These ranges were primarily derived from the 95% confidence intervals reported in previous studies. Sensitivity analyses therefore examined how the estimated reductions in deaths, injuries, and disability-adjusted life years (DALYs) changed when these lower and upper bounds were applied. This approach allowed us to capture the potential variation in outcomes associated with uncertainty in the effectiveness of the evaluated safety technologies [10,11,15].

## Results

In 2019, there were 18,489 road traffic deaths and over one million injuries, of which 63% of deaths and 48% of injuries occurred among vulnerable road users such as pedestrians, motorized two-wheelers, and bicyclists. Road traffic deaths and injuries caused 1,025,498 DALYs (Table 2). Vulnerable users were the most affected, accounting for 55% of DALYs. Almost half of all deaths in 2019 occurred in individuals aged 20–49 years, after which the number of deaths gradually decreased with age. There was a large gap between males and females, with most deaths between ages 15 and 84 occurring among males. However, this gap was smaller among younger (0–14 years) than older (85+ years) age groups. (Fig 2)

**Table 2.**
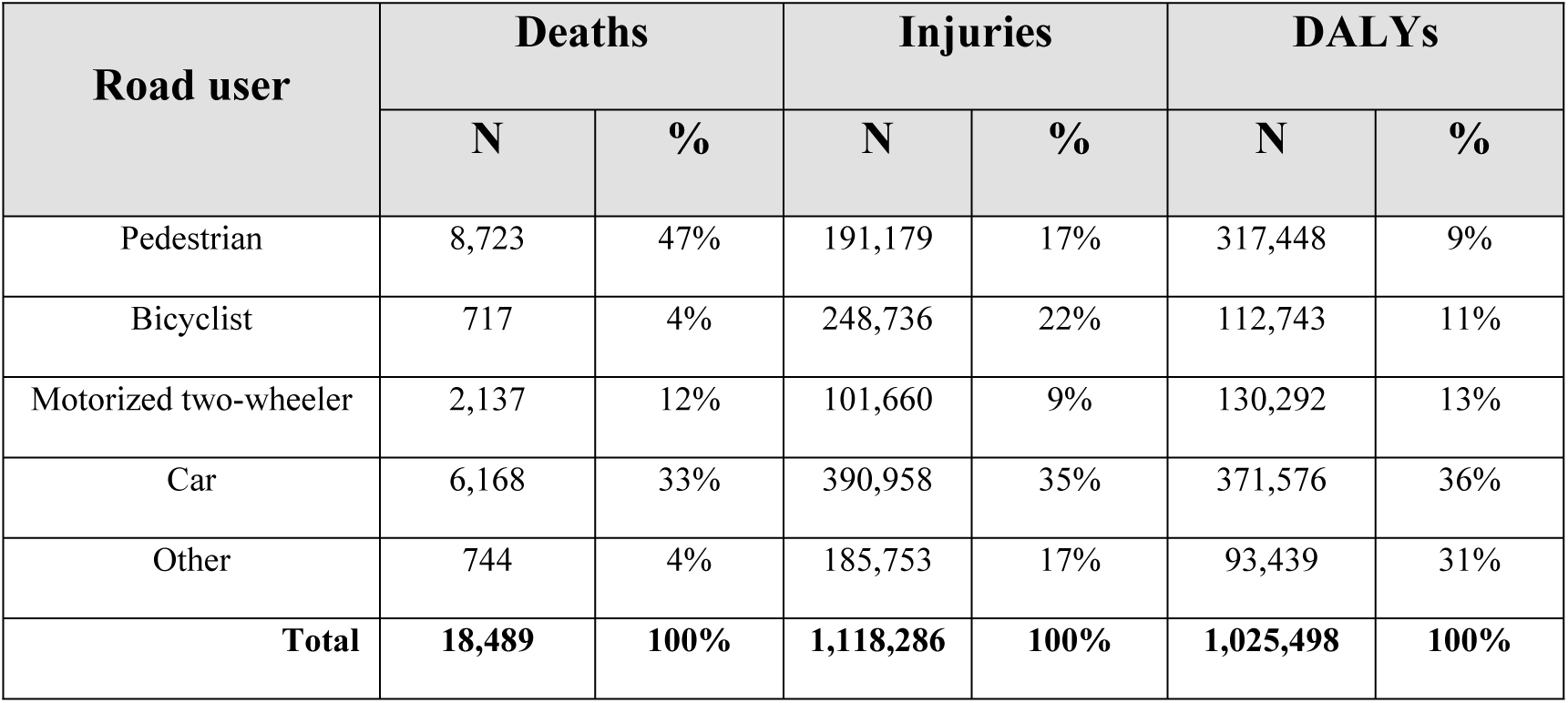
Descriptive results of the burden from road traffic injuries at baseline, 2019.

**Fig 2.**
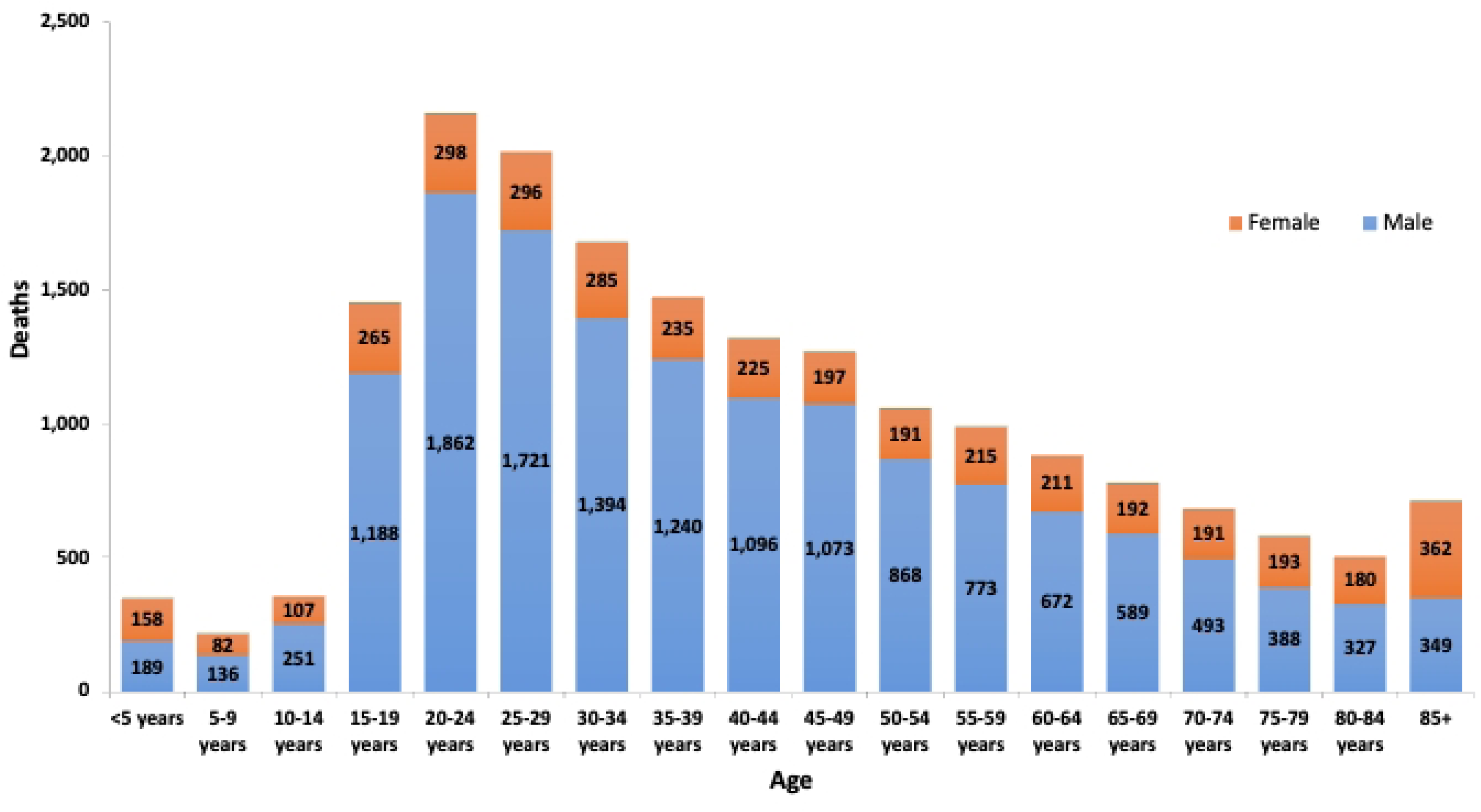
Road traffic deaths by age group and sex in 2019

### Scenario 1: Full compliance with 2022 legislation

Table 3 shows that full compliance with Mexico’s 2022 legislation is projected to yield substantial public health gains, reducing yearly road traffic mortality by approximately 18% (between 3,004 and 3,566 deaths), injuries by 16% (between 122,630 and 234,458 injuries), and DALYs by 18% (167,662 to 198,984 DALYs) compared to 2019. Fig 3 shows that the greatest benefits in this scenario are expected among car occupants, with reductions of 36% in deaths and injuries and 37% in DALYs, followed by motorcyclists with 20%, 36%, and 24% reductions in deaths, injuries, and DALYs, respectively, while pedestrians and cyclists experience a 7% decline in deaths. Both males and females are expected to benefit similarly, with an 18% reduction in deaths and DALYs (2,614 male and 707 female deaths prevented) (S5 Fig ***6***), and the largest absolute impact will occur among adults aged 20–64 years (2,460 deaths and 139,444 DALYs averted), followed by adolescents aged 15–19 years (306 deaths and 25,091 DALYs prevented) (S6 Fig ***7***).

**Table 3.** Reduction in deaths, injuries and DALYs in each scenario vs 2019 baseline.

|  | **Total** | **Averted number**<br>**(range from sensitivity**<br>**analysis)** | **Reduction percentage**<br>**(range from sensitivity**<br>**analysis)** |
| --- | --- | --- | --- |
| **Baseline deaths** | **18,489** |  |  |
| Scenario 1 | 15,168 | 3,321 (3,004-3,566) | 18.0 % (16.2-19.3) |
| Scenario 2 | 13,052 | 5,438 (4,972-6,551) | 29.4 % (26.9-35.4) |
| Scenario 3 | 10,985 | 7,505 (6,288-8,285) | 40.6 % (34.0-44.8) |
| **Baseline injuries** | **1,118,286** |  |  |
| Scenario 1 | 939,742 | 178,544 (122,630-234,458) | 16.0 % (11.0-21.0) |
| Scenario 2 | 814,415 | 303,871 (247,957-359,785) | 27.2 % (22.2-32.2) |
| Scenario 3 | 699,351 | 418,935 (360,943-472,772) | 37.5 % (32.3-42.3) |
| **Baseline DALYs** | **1,025,498** |  |  |
| Scenario 1 | 839,624 | 185,874 (167,662-198,984) | 18.1 % (16.3-19.4) |
| Scenario 2 | 721,139 | 304,359 (271,990-358,387) | 29.7 % (26.5-34.9) |
| Scenario 3 | 605,758 | 419,740 (350,767-462,187) | 40.9 % (34.2-45.1) |

Note: *Scenario 1* corresponds to full compliance with Mexico's 2022 regulation; *Scenario 2* includes scenario 1 and additional technologies in WP.29/WHO; and lastly, *Scenario 3* includes both previous scenarios and additional emerging technologies (ADAS).

**Fig 3.**
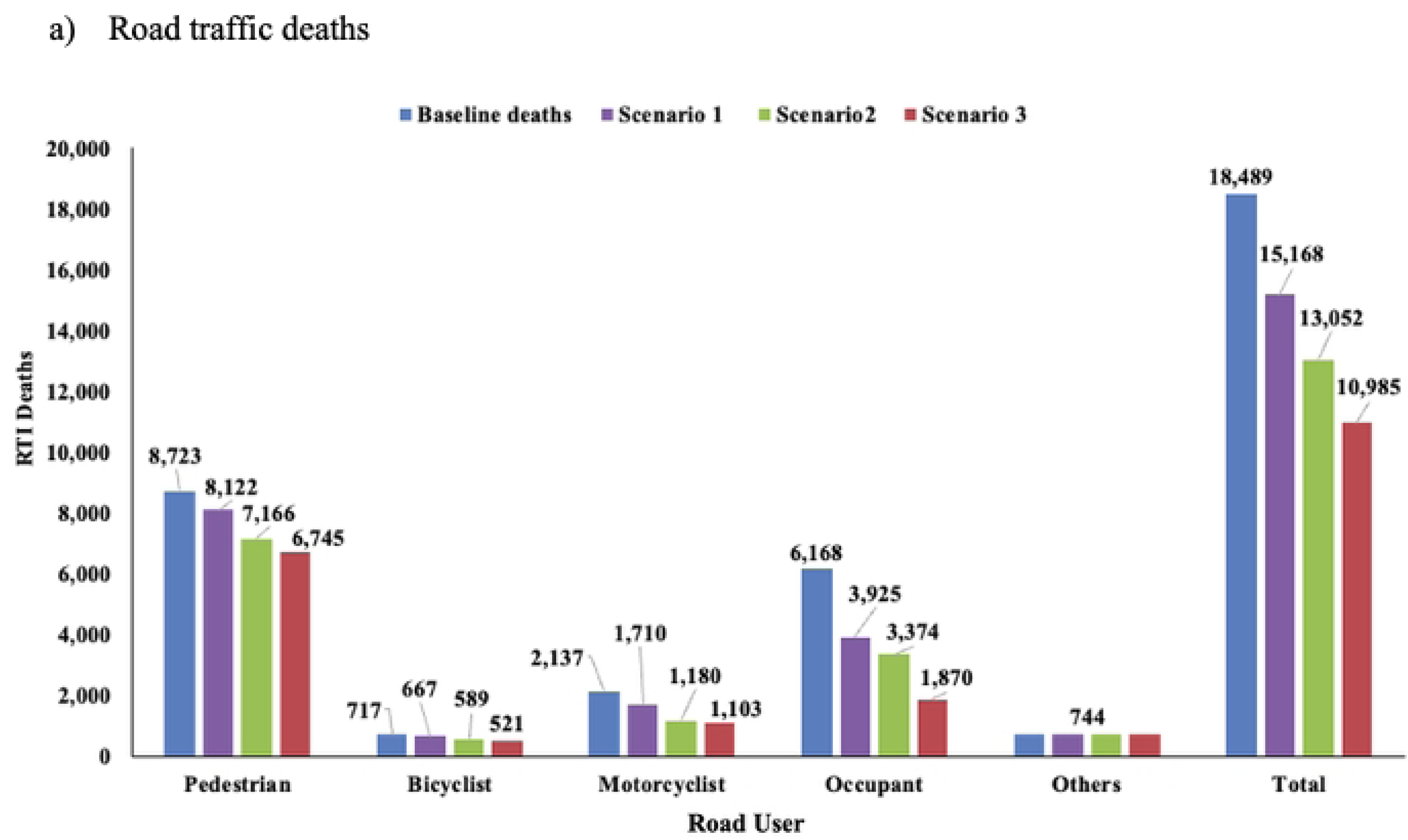
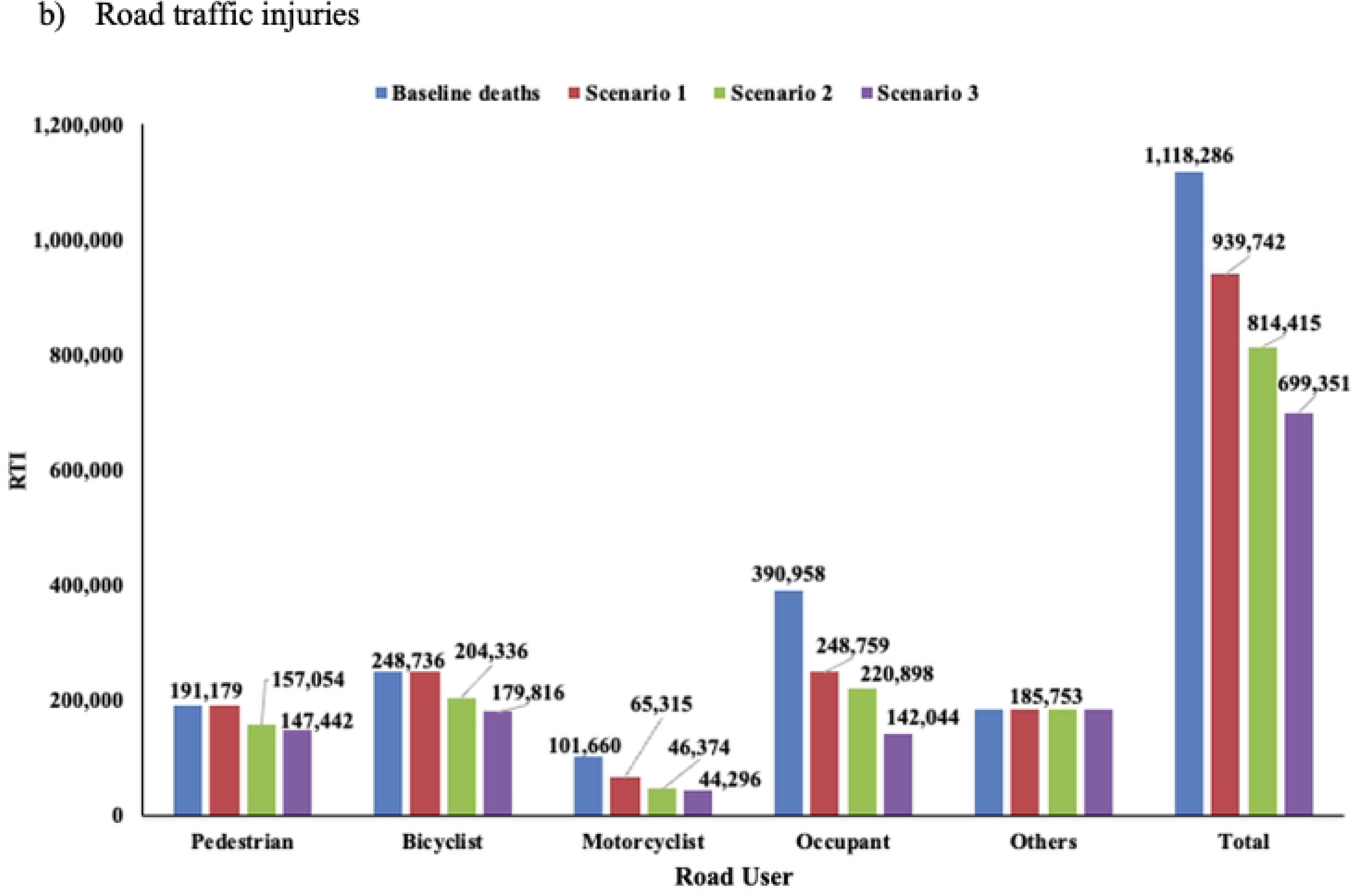
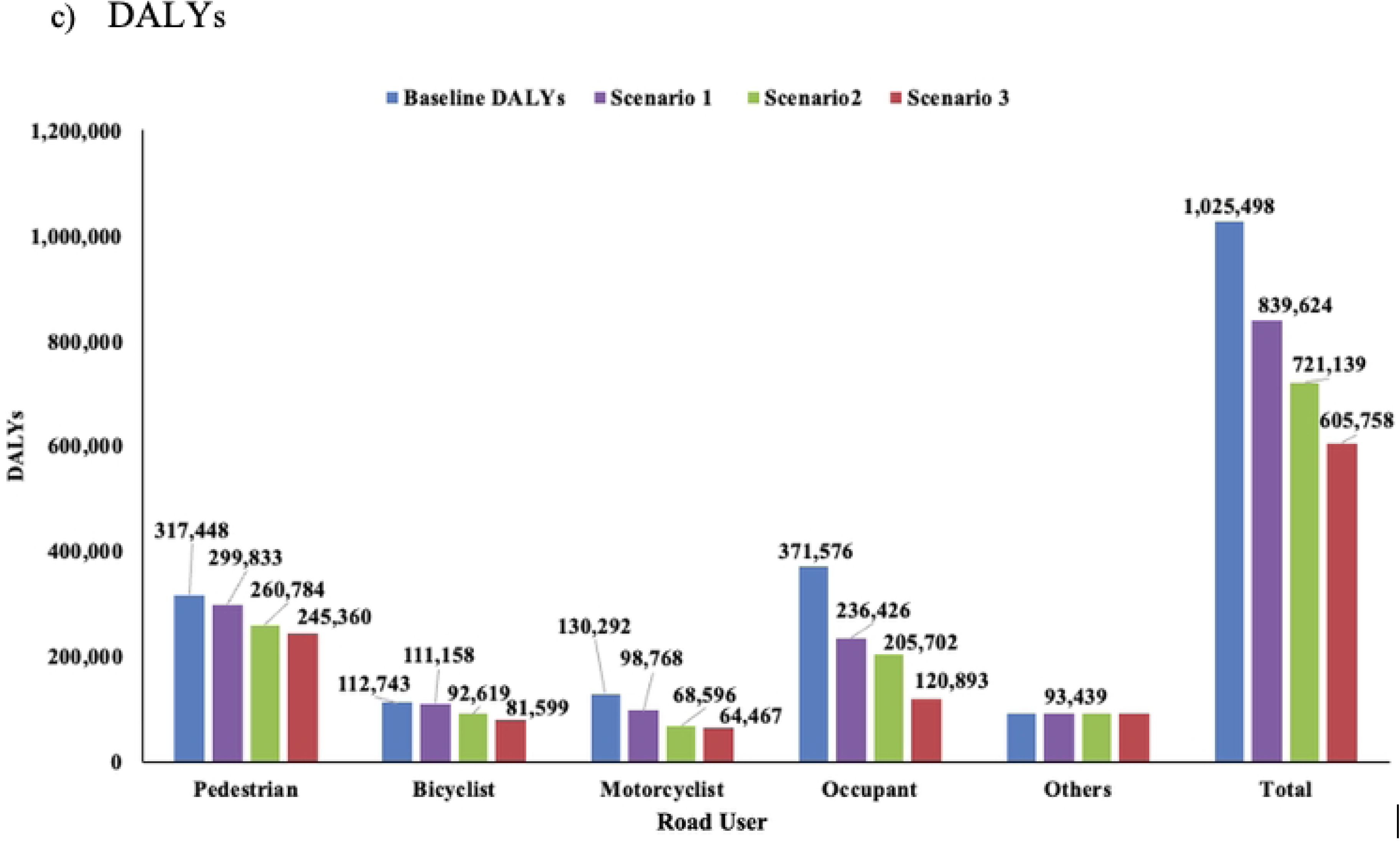
Summary results of all three scenarios compared to the 2019 baseline: total road traffic deaths, injuries, and DALYs by road user.

### Scenario 2: Expanding 2022 legislation to incorporate AEBS, crashworthiness, and ABS/ESC for motorcycles

Expanding current legislation to include technologies recommended by WP29, such as ABS/ESC for motorcycles, AEBS for rear-end collisions, and crashworthiness, is projected to amplify safety benefits, with estimated reductions of 29% in deaths (4,972 to 6,551 fewer deaths), 27% in injuries (247,957 to 359,785 fewer injuries), and 30% in DALYs (271,990 to 358,387 fewer DALYs) compared to 2019. Across road user types, motorcyclists would experience the greatest benefits, with 45% fewer deaths and 45–47% fewer DALYs, while pedestrians and cyclists could see an 18% decline in fatalities. Motorcyclist injuries could decrease by 54%, followed by 43% among car occupants and 18% among pedestrians. By sex, both males and females would experience similar relative reductions, around 29% in deaths and 30% in DALYs (S5 Fig ***6***). While the 15–19-year-old group is projected to have the greatest proportional benefit (34% fewer deaths), adults aged 20–64 years would account for the largest absolute reduction, with over 3,900 deaths and 223,423 DALYs prevented (S6 Fig ***7***).

### Scenario 3: Previous scenarios, ADAS, and expanding AEBS use

Further expanding Scenario 2 to include emerging vehicle safety technologies, such as AEBS for multiple crash configurations, AEBS for motorcycles, and advanced driver-assistance systems (ADAS), is projected to yield the greatest benefits overall, reducing road traffic deaths by 41% (6,288 to 8,285 fewer deaths), injuries by 37% (360,943 to 472,772 fewer injuries), and DALYs by 41% (350,767 to 462,187 fewer DALYs). A broader application of AEBS (for car-to-car rear-end and intersection crashes, car-to-pedestrian crashes, and car-to-bicycle crashes) alone could result in up to 13% fewer injuries and 15% fewer deaths and DALYs. Additionally, newer technologies, like Lane Departure Warning (LDW) and Lane Keeping Assist (LKA), could result in a reduction of 3 and 8% in deaths and injuries and 4 and 10% in DALYs, respectively. Across road user groups, car occupants would achieve the largest gains, with reductions of 70% in deaths, 64% in injuries, and 67% in DALYs, followed by motorcyclists (48%, 56%, and 51%, respectively). Cyclists and pedestrians would also benefit, with deaths decreasing by 27% and 23%, respectively (see Fig 3). By sex, reductions would be consistent, with 40% fewer male deaths (5,916) and 41% fewer female deaths (1,588), each experiencing a 41% decrease in DALYs (S5 Fig ***6***).

Adolescents would have the greatest proportional improvement, with 46% fewer deaths and 42% fewer DALYs, while adults aged 20–64 years would have the largest absolute impact, preventing approximately 5,466 deaths and 307,392 DALYs. (S6 Fig ***7***).

## Discussion

Our study shows that complete implementation of the 2022 legislation could lead to a yearly reduction in road traffic deaths (−18%), injuries (−16%), and DALYs (−18%) compared with the baseline year of 2019. We also show that upgrading the current legislation to include crashworthiness, ABS/ESC for motorcycles, and AEBS for rear-end car-to-car collisions could have important additional benefits, especially for vulnerable road users. This upgrade is in line with international recommendations for vehicle safety [13,48]. Our study provides the most up-to-date and robust estimates for the potential impact of vehicle safety legislation in Mexico, improving previous estimates [10] by using updated, country-specific data and analyzing a wider set of safety technologies, including motorcycle helmets and other safety standards for motorcycles, child restraint systems, and emerging technologies.

The approval and adoption of NOM-194 in 2022 is expected to have a large positive public health impact, as show in our study. Although reaching 100% conformity in the vehicle fleet will take time, NOM-194 provides clear procedures for implementation and evaluation, requiring manufacturers and importers to undergo formal conformity assessments by accredited inspection units that verify the presence and functionality of safety technologies before vehicles enter the market [22]. Uptake of safer cars among consumers could be accelerated by supporting independent consumer information programs such as New Car Assessment Programs (NCAPs). NCAP programs provide independent, transparent, comparable safety ratings that inform consumers and large fleet managers, and it encourages manufacturers to compete to achieve better ratings in the NCAP [6]. Another emerging strategy to inform consumers and promote market uptake of safer cars is vehicle safety labeling. This strategy could support the effective implementation of regulations, such as Mexico’s NOM-194, helping to accelerate its public health benefits.

Our study also shows that although the public health impact of the current legislation is very significant especially for car occupants, it is not protecting vulnerable road users because it excluded key technologies (crashworthiness, ABS/ESC for motorcycles and AEBS). AEBS can detect an imminent collision and automatically apply the brakes to prevent or mitigate impact [49]; pedestrian-friendly vehicle design integrates both active systems that avoid crashes and passive structures that reduce trauma when collisions occur [50]; and motorcycle ABS and ESC help riders maintain stability during sudden braking [51]. Enhancing NOM-194 to include these technologies could result in a 29% reduction in deaths, 27% in injuries, and 30% in DALYs, with vulnerable road users benefiting the most. Our estimates are consistent with findings from other studies using similar modeling approaches [7,14,15]. Although increased production costs are often cited as barriers to adoption, multiple analyses have shown that these technologies benefit from economies of scale and are highly cost-effective in preventing fatalities and reducing injury severity [15,52,53].

Technologies such as LDW and LKA (as modeled in scenario 3) are relatively new in Mexico and are present in only a small portion of the vehicle fleet. These systems are not yet mandatory under NOM-194, and additional research is needed to document their benefits and real-world impact on preventing road traffic injuries, with the goal of supporting their inclusion in future updates of the standard. Advanced driver assistance systems (ADAS) are expanding rapidly in markets such as the United States and Europe, where their safety benefits are increasingly recognized [54,55]. As vehicle safety technologies continue to evolve, it is essential that Mexico’s NOM-194 be regularly updated to remain aligned with technological advances and incorporate all relevant systems. Because technology advances more quickly than legislation, Mexico requires a framework for regulatory agility, ensuring that as ADAS and other innovations mature globally, they are integrated into local standards without the typical decade-long delays

Our findings highlight the public health benefits that can be achieved by improving vehicle safety standards in Mexico. Vehicle safety technologies must be complemented with other road safety strategies to achieve Decade of Action safety goals. Consistent with the Safe System approach, complementary strategies, such as designing forgiving roadway environments, promoting safe and responsible road use, managing speeds through context-appropriate design and enforcement, and strengthening post-crash care, are essential to maximize impact. When combined, improved vehicle safety and coordinated system-wide interventions can help Mexico move decisively toward the Decade of Action goal of reducing road traffic injuries and deaths by at leats 50%.

Our analysis has several limitations. First, the GBD data tends to overestimate the number of injuries in Mexico [26,56]. This may result in an overestimation of the absolute number of injuries prevented as well as the DALYs estimation; this, however, should not affect relative reductions. Second, the estimate of vehicle safety technology penetration has some limitations. EPC data covered brands/models for approximately 70% of vehicles sold in Mexico, focusing mainly on top-selling models in 2019. While the exclusion of high-end models may lead to a slight underestimation of overall technology prevalence, the 70% market coverage analyzed represents the vehicles most accessible to the general public. From a public health perspective, this segment of vehicles is the primary driver of road traffic injury statistics. Thus, we believe that our estimate for penetration in 2019 is robust, as it is based on the measured prevalence of each technology and considers the estimated prevalence of vehicle safety technologies among older models still in circulation. Third, RRs estimates derived from evaluations in high-income countries may not be directly applicable to the Mexican context. Differences in vehicle crashworthiness, maintenance practices, and the correct use of seat belts and child restraints, as well as weaker speed enforcement, road and infrastructure conditions, and less stringent driver licensing requirements, may substantially modify their real-world impact. Further on this point, although we have carefully applied risk reductions to recognize that technologies do not work independently, there may be some overlap which is not considered, especially in scenario 3. The sensitivity analysis is important to understand the range of potential effect of the scenarios. Finally, the model assumes correct functioning and user acceptance of technologies; yet, drivers may disable or override [57], advanced assistance systems, and infrastructure limitations may reduce real-world effectiveness.

## Conclusion

Vehicle safety technologies save lives and prevent injuries and disability. In Mexico, it is essential to ensure the effective implementation of NOM-194 and to accelerate market uptake of safer cars. Further, our study suggest that NOM-194 should be revised in the short term to include technologies such as ABS and ESC for motorcycles, AEBS, and pedestrian protection design to protect vulnerable road users. In the longer term, Mexico should evaluate the phased integration of technologies and advanced systems such as LDW and LKA.

## Data Availability

No data was generated by this study. The following existing data sources were used: Instituto Nacional de Estadística y Geografía (INEGI). Estadísticas de Defunciones Registradas (EDR) [Internet]. 2019 [cited 2025 Jun 23]. Available from: https://www.inegi.org.mx/programas/edr/ Consejo Nacional de Población. Consejo Nacional de Población. Proyecciones de la Población de México y de las Entidades Federativas, 2016-2050 [Internet]. [cited 2024 Sep 8]. Available from: https://datos.gob.mx/busca/dataset/proyecciones-de-la-poblacion-de-mexico-y-de-las-entidades-federativas-2020-2070

https://www.inegi.org.mx/programas/edr/

## Acknowledgments

This study was funded by the Global Road Safety Partnership (GRSP) grant MEXXX-VSRD22-2057. We would also like to acknowledge the support of the Global Health Advocacy Incubator and of our fellow GRSP grantees: El Poder del Consumidor, CENTRICO, Bicitekas, ITDP, Polithink, Instituto Sur, Salud Justa, WRI and the Mexican Red Cross. Further, we would like to acknowledge Edith Reyes and Alma Delia Sánchez for their administrative support, Alma Lizeth Alvarado, Alex Quistberg, Jessica Uruchima, Goro Yamada, Alvaro Guzman and Valentina Ochoa.

## Supporting information

**S1 Fig 4. Implementation of technologies by scenario**

**S2 Table 4. International Classification of Diseases 10th Revision (ICD-10) included in analysis - Transport Accidents**

**S3 Table 5. Safety technology prevalence in Mexico as of 2019 and their associated relative risks of injury and death.**

**S4 Fig 5. Comparison results of the reduction in road traffic deaths, injuries and DALYs by safety technology among the three scenarios.**

**S5 Fig 6. Summary results of each scenario compared to baseline: Total road traffic deaths and DALYs by sex, 2019 baseline vs all three scenarios.**

**S6 Fig 7. Summary results of each scenario compared to baseline: Total road traffic deaths and DALYs reduction by age group in 2019 baseline vs the three scenarios.**

